# Psychological safety, hierarchy, and other issues in operating room debriefing: reflexive thematic analysis of interviews from the frontline

**DOI:** 10.1101/2022.09.23.22280268

**Authors:** Canice McElroy, Emma Skegg, Mercedes Mudgway, Ngaire Murray, Linda Holmes, Jennifer Weller, James Hamill

## Abstract

**Background:** Debriefing is a team discussion in a constructive, supportive environment. Barriers exist to consistent, effective, operative team debriefing.

**Aim:** To identify barriers to debriefing and their potential solutions as articulated by operating room personnel.

**Design:** Qualitative study.

**Methods:** Between December 2021 and February 2022 we interviewed operating room workers in a tertiary children’s hospital. We used purposive sampling to interview a variety of professions and specialties who work in the operating room environment. Interviews were audio-recorded, transcribed, and coded. The qualitative approach was reflexive thematic analysis with the theoretical framework was critical realism.

**Results:** Interviews were analysed from 40 operating room staff: 14 nurses,7 anaesthetic technicians, 7 anaesthetists, and 12 surgeons; 25 (62%) were female. The five key themes were: 1) “commitment to learning” – healthcare workers are committed to teamwork, quality improvement, and teamwork; 2) “it’s a safe space” – psychological safety is a pre-requisite for, and is enhanced by, debriefing; “natural born leader” – the value of leadership and also the limitations caused by psychological constructs about what and who is a leader; 4) “space-time” – finding time to debrief after routine operations and after critical events; and 5) “doing the basics well” – debriefing needs structure without being over-complicated.

**Conclusion:** Psychological safety is both a prerequisite for and a product of debriefing. Leadership, if viewed as a collective responsibility, could help break down power structures. Given the results of this study and evidence in the literature, it is likely that routine debriefing, if well done, will improve psychological safety, facilitate team learning, reduce errors, and improve patient safety.

**Relevance to Clinical Practice:** Debriefing is challenging to perform, requires leadership and training, but is worth the effort. Nurses can take a leading role in promoting routine debriefing in healthcare.

**What does this paper contribute to the wider global clinical community?:**

- Debriefing, if done well, promotes teamwork, psychological safety in the workplace, quality and safety, and organisational learning.
- Setting up an effective debriefing programme is challenging but worth the effort.
- Debriefing requires collaboration and nurses are well placed to be leaders in debriefing policy design and to lead multidisciplinary debriefs.

## Introduction & Background

Surgery is a high-risk undertaking in which professionals from a variety of specialties work together on a time-critical operation in which safety is of the utmost importance. Similar to teams in aviation, motor racing and sport, surgical teams can be described as action teams. Where surgical teams differ is the diversity of roles within the group: nurses, anaesthetists, technicians, surgeons, from students to seniors. To perform effectively, team members must feel safe about sharing observations and opinions with the rest of the team (A. C. Edmondson, 2019); however, diverse surgical teams are often beset by intergroup rivalries based on profession, gender, and seniority. The resulting power dichotomies inhibit effective teamwork.

One way to counter this problem with surgical teams may be through regular team debriefing. Debriefing is a process that allows individuals to discuss team performance in a constructive, supportive environment (Fanning & Gaba, 2007). Debriefing has been linked to improved performance, teamwork and communication, and can assist with error identification (Zuckerman et al., 2012).

Debriefing is one component of the World Health Organisation Surgical Safety Checklist (SSC). Initially a three-point checklist involving a sign-in, time-out, and sign-out (Haynes et al., 2009), the SSC was expanded to include five steps by adding a briefing discussion at the beginning of the operating list, and a debriefing at the end of the list (Vickers, 2011). The sign-in, time-out, and sign-out are now performed with a high level of compliance in operating rooms throughout the world (Abbott et al., 2018). In our tertiary paediatric hospital, compliance with the SSC is 100% but compliance with debriefing (which is not mandated at the present time) is <10% (personal communication, Starship Children’s Hospital Operating Rooms, 2022).

Other authors have also found debriefing challenging to implement (Brindle et al., 2018), possibly because it involves a coordinated team discussion rather than a simple checklist, or for reasons related to power dynamics. Debriefing has roots in experiential learning theory (A. Y. Kolb & Kolb, 2006) and is essential to team learning (D. A. Kolb, 2015; Kolbe et al., 2020; Vashdi et al., 2013). Given that debriefing is central to other fields including medical simulation (Fanning & Gaba, 2007; Raemer et al., 2011), the aviation industry (Mavin et al., 2018), the space industry (Landon et al., 2018), and the military (Crane et al., 2006), we appear to be missing out on debriefing’s benefits in clinical healthcare.

Our surgical unit consists of seven operating rooms within a tertiary referral paediatric hospital providing cardiac, ear nose and throat (ENT), orthopaedic, thoracic, urology, and general paediatric surgical services. We introduced debriefing into our operating rooms in 2017. Despite the positive feedback, debriefing did not become routine practice throughout our operating rooms, being performed in only a minority of operating lists. Given the potential value of debriefing for surgical teams but the challenges of putting it into practice, we were interested in the views of our operating room staff on the how, why, when and where to debrief.

The purpose of this study was to gain insights from frontline workers on how to set up an effective debriefing policy for our operating room suite, to explore operating room workers’ experiences with debriefing, and to critically reflect on the meaning of debriefing to psychological safety, hierarchy, and teamwork.

## Methods

### Ethics and Trial Registration

This study was registered with the institutional research office and received ethical approval from the Auckland Health Research Ethics Committee (#3228).

### Context

The setting was the operating room of a children’s hospital. Operating room workers had some experience with debriefing since it was informally introduced in 2017. A simple debriefing structure was developed which consisted of three phases: 1) introductions and ground rules (confidentiality, respect, share the air-time); 2) discussion (what went well, what could have gone better); and 3) take home actions. This structure was used mostly in paediatric surgery and in ear, nose and throat operating lists. Paediatric cardiac surgery regularly debriefed all operations using an extended checklist structure. Apart from cardiac surgery, debriefings were performed inconsistently and were not part of the operating room culture.

### Researcher Characteristics & Reflexivity

Researchers brought together a mix of experience and fresh eyes on the subject. Two researchers were senior nurses who had been instrumental in introducing debriefing in 2017. At the time of the study, one was a senior operating room nurse (LH) and the other was the nurse manager of perioperative services at Starship Children’s Hospital (NM). One researcher was a psychology masters graduate with background in neuroscience (MM). Two researchers were medical students (CM and ES). Two researchers were doctors: JW is an anaesthetist with extensive experience in simulation training, debriefing and multidisciplinary teamwork; and JH is a paediatric surgeon with experience in debriefing in simulation training and in operating room. CM and ES performed all interviews; as students, they came from a more independent stance than some of the other researchers who were co-workers and colleagues of the participants. Theme development and critical interpretation was performed by a student (CM), psychologist (MM), and operating room worker (JH), bringing a variety of perspectives on the topics.

### Qualitative Approach & Research Paradigm

The qualitative methodology used for this research was reflexive thematic analysis (Braun & Clarke, 2022a). The theoretical framework was one of critical realism (Bhaskar, 2013). According to Pilgrim (Pilgrim, 2014), in critical realism “both causes and meanings are given due weight in social scientific enquiries and the entwined relationship between facts and values emphasised.” According to Yucel (Yucel, 2018), critical realism “avoids many of the problems of positivism and social constructivism by finding a middle ground between them.” Operating room personnel work in a technological environment. We wanted to “give voice” to what people told us in the interviews while recognising that how we understand these “truths” is focused through the psychosocial lens of the interviewers and the interviewees. As operating teams, we deal in reality – “life and limb” – so reality matters; however, we recognise that social constructs determine how team members relate to each other, hence the reason for choosing a critical realism paradigm. Our purpose was to shed light on the barriers to debriefing and to identify factors that might facilitate routine debriefing. Critical realism and reflexive thematic analysis also allowed us to recognise the transitive nature of the research where the researchers are part of the study.

### Sampling Strategy & Data Collection

The inclusion criteria were operating room staff including nurses, anaesthetic technicians, anaesthetists, and surgeons, who worked regularly in the children’s operating suit. Excluded were students and other temporary visitors to the operating rooms. The sampling strategy was purposive, with care taken to interview a range of professions and specialty groups. The sample size was determined pragmatically by the number of interviews able to be performed over the study period, December 2021 to February 2022, but also guided reflexively as recommended by Braun and Clarke (Braun & Clarke, 2021a, 2022a), and Malterud’s “information power” concept helped guide the decision as to when to stop recruiting (Malterud et al., 2016). Our considerations in determining the information power of our sample were: 1) the aims of this study was quite broad, favouring a larger sample; 2) the specificity of the sample was relatively broad, favouring a larger sample; 3) there was no established theory, favouring a larger sample; 4) the quality of the dialogue in our interviews was strong, favouring a smaller sample; and 5) the analysis was across cases, favouring a larger sample.

Operating room staff were made aware of the project through presentations at meetings. Two researchers (CM and ES) invited participants to take part in an interview. Participants gave written informed consent. Interviews were conducted between December 2021 and February 2022. All interviews took place in person in a private space using a semi-structured interview guide (available in Appendix 1 in supplementary material). The interview guide was developed by the researchers to cover the items of interest based on our experience with surgical debriefing (e.g., issues of timing and structure). Limited testing of the interview guide was undertaken by mock interviews between three researchers. Interviews began with making personal connections and explaining the study, then proceeded with open-ended questions with an emphasis on letting the interviewee tell their story and do most of the talking. Interviews were audio recorded using a mobile device and transcribed verbatim. Participants received a copy of their transcript for editing and approval. Transcripts were de-identified and labeled with a random number generated from the random sequence generator service of random.org (Haahr, 2023).

### Data Processing & Analysis

We followed the process of reflexive thematic analysis described by Braun and Clarke (Braun & Clarke, 2022b). Briefly, coding and theme development were recursive processes involving immersion in data, review of relevant literature, and deep reflection. Immersion in the dataset was by transcribing (performed by CM and ES), reading, and re-reading interview transcripts. Two researchers (CM and JH) coded the data separately and met regularly to discuss and mould themes. The online version of Taguette (Rampin & Rampin, 2021) was used as an aid to coding. Taguette is an open source tool used to import transcripts, highlight and tag quotes. We drew mind maps to help visualise the diversity and connectedness of themes as they developed. We took both a semantic and latent approach to coding, meaning that some codes came directly from what interviewees said (semantic) while we developed other codes based on our understanding of teamwork and psychological safety (latent). While some of our analysis was inductive (coming straight from the data) much was deductive (our interpretation of the data as it related to debriefing in keeping with the “critical” aspect of the critical realism paradigm). Through a process of team discussion and reflection, we refined codes into initial themes. Mapping themes graphically helped demonstrate connections and interactions between initial themes. Through a process of reflection, discussion, re-reading transcripts, reflecting on initial themes, refined and renaming, we developed the main themes of the research.

### Techniques to Enhance Trustworthiness

To enhance trustworthiness of this report, we took a lead from Braun and Clarke’s guidance on quality and reporting (Braun & Clarke, 2021b) and from the guidelines for publication of qualitative research by Elliot et al. (Elliott et al., 1999). First, we applied Braun and Clarke’s 15-point checklist of criteria for good thematic analysis to our study (Braun & Clarke, 2006) as shown in Appendix 2 in supplementary material. Next, we used Nelson’s conceptual depth scale to self-evaluate robustness of our conceptual categories (Nelson, 2017) as shown in Appendix 3 in supplementary material. We used Braun and Clarke’s 20 questions for evaluating thematic analysis manuscripts for publication (Braun & Clarke, 2021b) as shown in Appendix 4 in supplementary material. The present report follows the Standards for Reporting Qualitative Research (O’Brien et al., 2014) as shown in Appendix 5 in supplementary material.

## Results

Of 41 interviews conducted, one recording failed leaving 40 interviews to be transcribed and analysed. Interviewees were predominantly female (n = 26, 65%). Interviewee characteristics are shown in Table 1. Each interview lasted approximately 10–20 minutes. Interview transcripts amounted to 84748 words, an average of over 2,100 words per transcript.

**Table 1.** Interviewee characteristics.

|  | n = 40 |
| --- | --- |
| Gender |  |
| Female | 25 |
| Male | 15 |
| Profession |  |
| Nursing | 14 |
| Anaesthesia | 7 |
| Anaesthetic technician | 7 |
| Surgeon | 12 |
| Surgical specialty |  |
| Paediatric surgery | 7 |
| Orthopaedic surgery | 2 |
| Cardiac surgery | 2 |
| ENT | 1 |

A map of initial themes is presented in Fig. S1 in supplementary material. Through a process of reflection, discussion, and re-reading transcripts, we refined the initial themes to five final themes. The development of these themes is shown in Fig. 1. A final table of themes is presented in Table 2. In the following paragraphs, we illustrate each theme with selected quotations from the interviews.

**Table 2.** Themes and description of each theme.

| Theme | Explanation |
| --- | --- |
| Committed to learning | Health workers are committed to learning, quality improvement, and to the performance of the organisation. |
| It's a safe space | Debriefing promotes psychological safety, psychological safety is needed in debriefs, and constructs around power dynamics in teams. |
| Natural born leader | Who should lead debriefs and constructs of the meaning of leadership in teams. |
| Space-time | Practicalities on when and where to debrief, the problem of people needing to leave, and constructs on the meaning of time. |
| Doing the basics well | Realist reflection on the right amount of structure in performing debriefs and on training how to debrief. |

**Figure 1.**
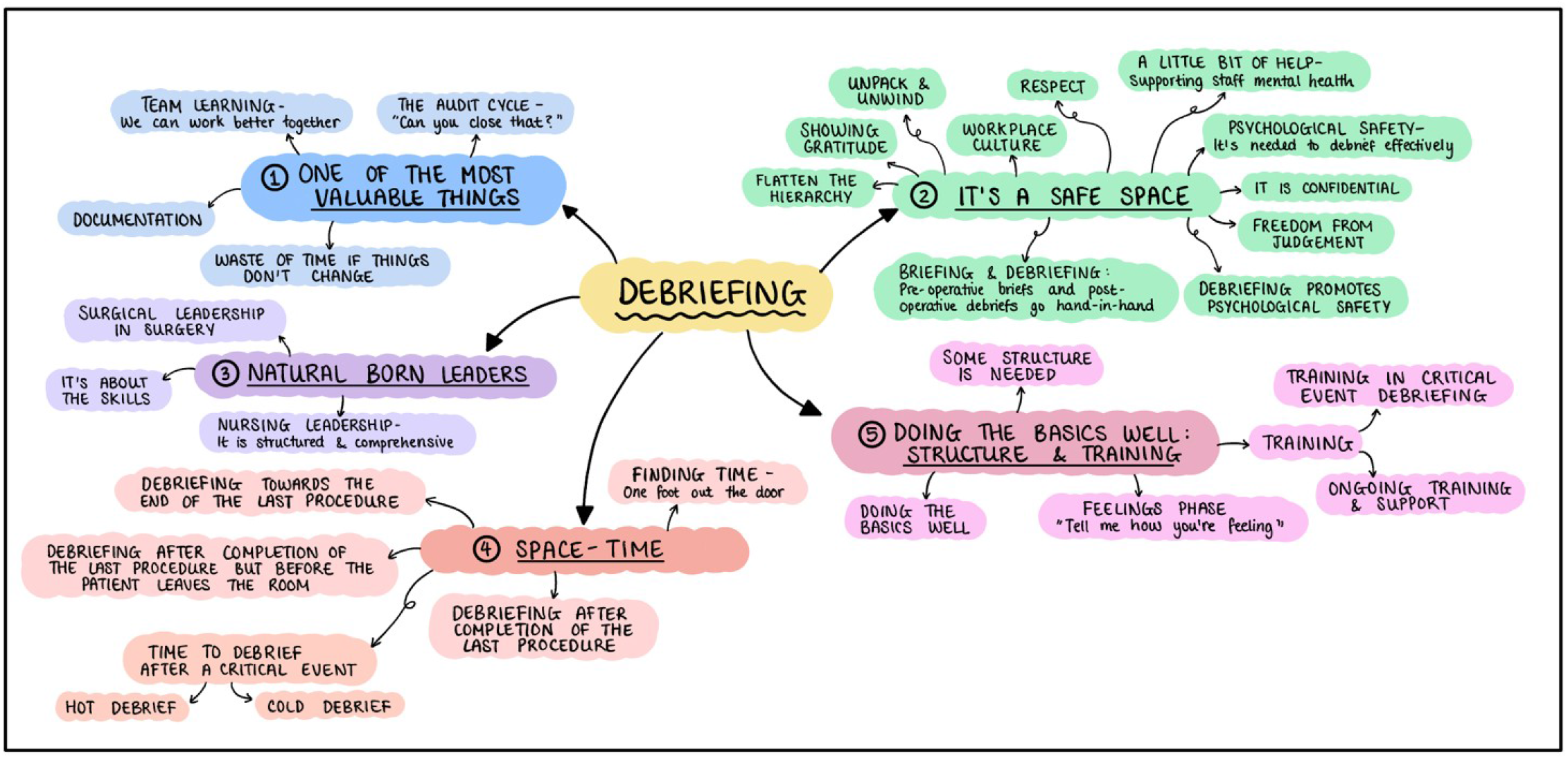
Mind map of themes developed from the study.

### Theme 1. Committed to learning

A strong theme was how committed frontline workers are to learning and quality improvement. Learning was seen as one of the most important things we do in healthcare and debriefs were seen as a vehicle for learning.

> “It’s just a way to kind of come together with the team at the end of the day and talk about any issues and try and think of possible solutions, so that when we do this, when we do the list again, we can work better and not have those issues.” (Participant 10)
>
> “Sometimes there’s quite good key learning points and from theatre, well if every theatre did a decent debrief at the end of the day, I think that there might be things that might come up consistently, repeatedly, that actually we can learn from – learning debriefs…” (Participant 36)
>
> “Is there anything that we can learn from this, you know that’s almost the most important thing in modern medicine and healthcare, isn’t it? You know it’s, ‘what can we do better next time so that we can avoid this situation?’” (Participant 4)

It was striking that staff are not just committed to individual and team learning, they want to see organisational change too. They wanted debriefs to be incorporated into a quality improvement process so that issues raised were dealt with at an organisational level. If debriefs did not lead to quality improvement, then they were seen as a waste of time. Conversely, if debriefs lead to learning they were seen as immensely valuable.

> “It’s kind of like you just get stuck in Groundhog [Day] with the same – you’re not coming to a conclusion or fixing the problem; it’s just you’re continuously talking about it.” (Participant 26)
>
> “… although if there is any big issue, I think if there’s anything that needs improving, it could be good to record… otherwise it won’t get fixed.” (Participant 13)
>
> “Can you close that? Can you come back to us in two weeks to know how you progress?” (Participant 14)
>
> “I’ve found it absolutely one of the most valuable things that we’ve had instituted here.” (Participant 38)

### Theme 2. It’s a Safe Space

The importance of being psychologically safe in the team was a strong theme throughout the interviews. Psychological safety was promoted *by* debriefing, but also a pre-requisite *for* effective debriefing.

> “And then also just making sure that if someone does say something they’re not going to be shot down or they’re not going to be – yeah, just making sure it’s a safe space to do so.” (Participant 11)

#### Debriefing Promotes Psychological Safety

Interviewees felt that consistent debriefing would facilitate better communication, help flatten the hierarchy, and create a more inclusive work environment. Many participants recognised the importance of acknowledging positives during debriefs. Positive feedback is an effective way to encourage speaking up. Developing a structure that makes space for positives (“what went well”) was helpful. Several respondents commented that debriefing is a good way to unpack what had happened during the day, unwind a bit, and to go home leaving work at work, which would clearly carry co-benefits for mental health and resilience. Debriefing critical events was also seen as essential for people’s mental health.

> “By doing briefing and debriefing it actually helps to break down barriers between different subgroups.” (Participant 2)
>
> “So sometime, often when the list is run really well, I think it’s really important to acknowledge that too, everybody to kind of be grateful to each other.” (Participant 36)
>
> “I think the debriefing actually finishes off the day and creates a good work environment and lets people kind of put work aside.” (Participant 36)
>
> “Everybody responds very differently to trauma and to, you know, critical incidents, and sometimes people just need that little bit of help and support to sort of express how they feel. Because otherwise you may end up losing these people because they just don’t want to do this job anymore.” (Participant 4)

#### Psychological Safety Facilitates Debriefing

Operating room staff needed to feel they were in a safe place to debrief effectively. Safety took various forms: assurance of confidentiality, a non-judgmental and respectful environment, and a flat hierarchy. People need to feel safe from being judged. Debriefs need to be conducted in a climate of respect. Creating an environment in which everyone, including inexperienced staff and students, can feel free to speak up is a challenge when there is an obvious hierarchy arrangement. There was an interplay between the tone set in the pre-operative briefing and that of the post-operative debriefing, and an interplay with the general culture of the operating room environment. Responses suggest that the attitude and approach to preoperative briefings impacts on the postoperative debriefings. Most interviewees felt that psychological safety is largely determined by broader operating room culture and attitudes in the workplace.

> “I think that starts from before the brief. So, I think having everyone there from the beginning, having an expectation that there’s a flattened hierarchy, we would welcome people speaking up, and that we use individual names, we introduce each other at the brief, we highlight how important each team member is.” (Participant 36)
>
> “In terms of like if people want to – anyone can say something, like it’s not a judgmental environment.” (Participant 26)
>
> “I think also that needs to be emphasised that it’s confidential.” (Participant 38)
>
> “I guess sometimes that’s where there might be issues with people not wanting to speak up if they feel that there’s a hierarchy.” (Participant 22)
>
> “It’s a longer-term thing… I think there’s an old-school way of practicing that you might not have even experienced that I experienced as a junior doctor – it’s fading out where there’s quite a lot of barriers to communication with senior people. And I think that’s improving, definitely, but I think it’s something that we generate every day through our interactions with other staff members. So I think it’s on the ward, it’s in pre-op, it’s in the clinic, it’s in the operating room, you just want to have an open, positive environment for discussion.” (Participant 28)

### Theme 3. Natural born leader

The theme of leadership considers interviewee perspectives on nominating a leader of the debrief session. Responses indicate that leadership is necessary to facilitate an efficient and effective debrief. In the following extract, an interviewee discussed options for leadership.

> “I think that’s a difficult one, actually. Because it could be different people on different days… it shouldn’t really be a set person every time. Because sometimes the surgeons change, sometimes the nurse in charge will pop out and then come back at the end. So really, there should be a natural leader that emerges, and sort of leads the debrief. But probably the person that needs to instigate it should be the nurse in charge, because otherwise surgeons might just forget to do it. So, it should be instigated by the nurse in charge, but then the natural leader should just do the chatting…” (Participant 18)

#### Surgical Leadership in Surgery

Many interviewees viewed senior doctors, the surgeon or the anaesthetist, as the main leaders in the OR.

> “In my experience, I think it’s best to have a surgeon or anaesthetist be onboard and leading it. I feel like they’re more of like a leader in the room and… in my experience people appear a lot more engaged when it’s an anaesthetist or a surgeon or someone quite senior in the room to be leading it.” (Participant 31)

#### Nursing Leadership is Structured and Comprehensive

Other interviewees felt that nurses were best suited to provide a comprehensive overview of operating room events and tended to follow a more structured and systematic approach.

> “The experience that generally I have is that the nurse that’s coordinating the list for the day runs it, and I think that allows for a lot more of an inclusive approach.” (Participant 20)

#### It’s About Skills

Many interviewees felt that anyone within the multidisciplinary operative team should be able to lead an operative team debrief. Responses suggest that an individual’s experience and communication skills may be more relevant to a leadership position than their title or specialty. Therefore, the leadership role can be flexible and negotiated.

> “I don’t think it should be any particular person, I don’t think. But I think the person needs to be able to lead it, so they need a sense of confidence, and they need to be able to understand when the most appropriate time is for debriefing… So, you have to be experienced enough to know – pick up on those sorts of things, but also subtle clues as well that maybe something is going on, it’s not quite the right time. So, you need a bit of experience I think to lead the debrief, and very much so, confidence. So, you need to be a little bit more experienced, but I don’t think necessarily it should be one person’s role.” (Participant 7)

### Theme 4. Space-time

There was little debate about the right place to debrief – consistently people found the practical place to be in the same operating room where the case(s) had been performed. Finding “space” in the sense of time in the day, however, was a challenge.

#### One Foot out the Door – Routine Debriefing

Some participants suggested that debriefing should occur whilst closing the skin of the final case. Interviewees felt that this approach would be the most achievable way to ensure that all team members are still present and engaged. At this point, the most challenging parts of the operative procedure have been completed, and the anaesthetic team is yet to begin waking up the patient. Responses suggest that during this time window, team members were still focused on the current session, which may be optimal for recalling learning points from the preceding list. Additionally, this time avoids the challenge of trying to bring team members back, which is practically challenging after the list is finished.

> “Because pretty much as soon as we start closing people start wandering off, and the surgeon has already got one foot out the door by the time the registrar or the fellow gets on to closing the skin layer. So I would have thought towards the end of the last operation would be the best time to catch everyone who was in that session without delaying people who want to get away at the end of the day.” (Participant 6)

Other participants felt that the ideal time to debrief is at the end of the list, after the patient has been taken to recovery. Responses suggest that this approach is likely to be associated with fewer distractions and result in greater focus on the debrief itself. Interviewees also felt that because the whole list is finished, the debrief would be able to cover all aspects of the day, including the closing of the final case. Responses revealed the main drawback to this approach is the challenge to get all team members back to the operating room for a debrief.

> “I really think it should be done at the very end, when the patient has left the room. Because, and everyone should be, you should sort of say at the end of the case. ‘we’re going to do a debrief, so can we all meet back here in ten minutes’. And then it just means that everyone’s mind is on that rather than on the patient. And it also makes it a bit more, sort of, real, if everyone’s just focusing on that rather than focusing on other things.” (Participant 18)

#### Time to Debrief After a Critical Event

Debriefing critical events was seen as valuable but, again, there was a timing debate. Some believed that critical event debriefs were best performed straight after the event (“hot”); others felt that critical event debriefs should wait until things settle down (“cold”).

> “If there’s something happened, if there’s something wrong, like baby died on the table or something happened like an emergency or crisis that happens on the table, it should be right after. Not tomorrow, not the next day. Yeah, like on the day when everyone’s still around and you don’t have to look for them.” (Participant 41)
>
> “It should probably be dealt with a little bit later in a setting where everybody feels safe. And preferably the timing, depending on the personalities and how well you know the people, to be able to give a bit of time for people to be able to process it after a case, personally.” (Participant 5)

#### All the Time

One facet of the collective effort theme is that debriefing should be done consistently across all teams. This creates a universal standard of practice. Interviewees felt that collective support would be necessary to facilitate this cultural change.

> “The more they see people doing it… the more they feel that they can contribute. Because they’ve seen it before, it becomes more familiar, more routine, and they realise that there’s no negative repercussions for it.” (Participant 33)
>
> “It actually breaks down those barriers, and people feel that they can ask those questions… by doing them consistently is actually how we break down the barriers as well.” (Participant 2)

### Theme 5. Doing the Basics Well

Interviewees felt that debriefing techniques should be kept simple but implemented in an effective and efficient manner. There was clearly a need for some framework in which to construct a purposeful conversation. One aspect of structure was the “reactions” or feelings phase which commonly occurs near the start of simulation debriefs.

> “Try to keep doing the basics well. We’re not overcomplicating too much stuff; I think that would be the way to go.” (Participant 14)
>
> “I think it needs to be structured because otherwise it descends into waffle and stories and anecdotes and opinions rather than actually exchanging information that is relevant to how the day went and to use our time efficiently and particularly at the end of the day… I do think that it’s better having some structure, and it being led by someone with a particular interest in getting some useful information.” (Participant 6)
>
> “It also gives people an opportunity to tell how they’re feeling although often people won’t necessarily say how they’re feeling, but I think that’s the thing of a good debrief is to make people feel comfortable to do that and I think it’s good to do it even when everything went smoothly and everything went fine.” (Participant 38)

#### No-one is Born Knowing how to do this

Some participants expressed the need for teaching and modelling of debriefing skills. Critical event debriefing would be easier if there was a culture of routine debriefs and if staff had training in how to facilitate a debrief. Other interviewees expressed that ongoing training and education would be useful for debrief leaders.

> “No one is born knowing how to do this, you actually do need to have some training.” (Participant 14)
>
> “I think that hot debriefs are very difficult, and you need to be very very careful that they’re done safely. I wonder if we could… have a format and have a culture of how those should be carried out and maybe some specific people that might champion doing the hot debriefs that I think everyone should probably have some training on doing it safely.” (Participant 36)
>
> “I think some form of support for the people that lead the debrief would be useful, as well as teaching of certain skills saying that you know like, when we do this, these are the aims, and let’s try to keep things running that way.” (Participant 14)

## Discussion

By giving voice to frontline workers, we have attempted to understand what is important when working within an operating room environment. The themes reflected a rich sense of the value of debriefing for learning, culture, and togetherness as a team. Debriefing’s influence on a culture of gratitude and psychological safety came through strongly, as did the contribution debriefing could make to team performance. Our research has established a foundation for various factors that relate to psychological safety including power hierarchies and perceptions of leadership.

### Committed to Learning

The commitment to quality, safety, and organisational learning by frontline staff is not always appreciated and organisations can miss opportunities to tap into the intellectual value of employees. The literature on commitment of healthcare workers to organisational learning is sparse. In a qualitative study of frontline healthcare workers in London, Lalani et al. (2020) found that “participants from across health and social care expressed their ambition to work collaboratively and maximise opportunities for formal and informal learning” (Lalani et al., 2020). Fathy Ahmed et al. (2023) demonstrated a strong positive correlation between goal orientation in staff nurses (commitment to learning and performance) and organisational learning culture (dialogue, team learning, collaboration, empowerment, etc.) and recommended that nurses are empowered to participate in decision making, problem solving, and in achieving organisational goals (Fathy Ahmed et al., 2023). Our research showed that people working in theatre are committed to learning and improvement. Debriefs were seen as a vehicle for individual and team learning, with the potential to contribute to organisational change. Participants saw debriefs as a way to identify key learning points and improve the quality of care. However, they wanted debriefs to be incorporated into a quality improvement process to ensure that issues raised were dealt with at an organisational level. If debriefs did not lead to quality improvement, then they were seen as a waste of time. Overall, committed staff in healthcare are eager to learn and improve, and organisations would do well to value their input and incorporate their suggestions into quality improvement processes.

### Psychological Safety

Our study uncovered a prominent theme concerning psychological safety in the theatre environment, which was viewed by participants as paramount for effective debriefing. Despite this, we found concerning evidence that some theatre staff members did not perceive themselves to be in a psychologically safe working environment.

According to Edmondson (2008), psychological safety ensures that individuals are not punished or humiliated for expressing ideas, questions, concerns, or mistakes. Our study reveals that creating an environment where individuals can be open and confident in group situations depends on the prevailing attitudes and culture in the workplace. Therefore, psychological safety is shaped by broader cultural norms within the organisation. Research shows that teams who welcome feedback have the healthiest work environment by supporting staff that report problems without fear of retribution and accountability (A. C. Edmondson, 2003). This creates an environment focused on proactively identifying and addressing actual and potential safety concerns, building a safer health system for patients, and increasing psychological safety for staff (Leonard et al., 2004).

To create trust, some institutions incorporate programmes that acknowledge or give positive recognition for reporting (Trossman, 2019). Positive recognition can be implemented by thanking individuals for speaking up and acknowledging why reporting is necessary, reinforcing trust between groups (Trossman, 2019).

Eliminating the fear of consequence is crucial to building trust and establishing that reporting will not negatively affect the staff member voicing their concerns or the member of staff involved in a report (Dekker, 2016). During our research, we found that people felt comfortable to speak up in a debrief about anything that had gone wrong during surgery. It was a process of team reflecting, rather than an outlet of blame on an individual, which coincided with an elimination of fear of consequence. This elimination of fear of consequence further builds on staff trust, ensures a positive feedback loop, and provides a learning opportunity from the event that has been reported (Dekker, 2016; Waring, 2005). Further, our research recognised the importance of acknowledging positives during debriefs. Positive feedback was seen as an effective way to encourage individuals to speak up and developed a structure that makes space for positives. Developing a structure that makes space for positive feedback was seen as an effective way to encourage individuals to speak up.

In a healthcare setting, psychological safety and just culture are essential to promote a safe and effective workplace culture. However, social power and perceived hierarchies in the workplace can impede the implementation of these principles (Vanstone & Grierson, 2022). The idea of power hierarchies in the medical field stems from an understanding of hierarchies that can present themselves in various areas, with different levels of authority given to individuals based on their perceived rank (Hughes & Salas, 2013). These hierarchies can start implicitly from the day students start their professional training and can reflect biases held from a young age (Gopal et al., 2021). Research has shown that these power hierarchies can diminish the effectiveness of critical work teams and team performance (Hughes & Salas, 2013). One of the primary ways that power operates in the workplace is through the establishment and maintenance of hierarchies (Vanstone & Grierson, 2022). Hierarchies are a well-recognised feature of the clinical environment, and research indicates that they can impede patient safety by contributing to a reluctance to speak up if a superior makes an error (Brennan & Davidson, 2019; Vanstone & Grierson, 2022). Furthermore, hierarchies can obstruct interprofessional teamwork and collaboration, leading to a hostile work environment that impacts individual and team psychological safety (A. Edmondson, 2008; McClintock & Fainstad, 2022).

However, hierarchies can also provide a template for expectations of interaction that generate and sustain the structure and stability necessary to enable the formation and efficiency of the team (Knight & Mehta, 2017; O’Shea et al., 2019; Vanstone & Grierson, 2022). To promote a just culture and psychological safety, it is crucial to recognise and address the negative effects of power hierarchies in the workplace. By doing so, healthcare professionals can foster an environment that values open communication, teamwork, and safety (Vanstone & Grierson, 2022).

While hierarchy can hinder processes, it can also be necessary for achieving a team goal. Those in higher hierarchical positions in a surgical environment can not only set the mood for the day but can also create a positive team environment and initiate or delegate team debriefing, which has been found to have a positive impact on teamwork and communication and break down barriers of hierarchy (Berenholtz et al., 2009; Paull DE et al., 2010; Robinson et al., 2010). Furthermore, hierarchy can be understood in terms of leadership, where leaders in healthcare organisations play a crucial role in promoting a culture of safety and just culture by addressing power imbalances and promoting open communication (A. C. Edmondson & Lei, 2014; Nembhard & Edmondson, 2006).

### Leadership

Our research showed that participants had varying perspectives on leadership in the context of medical debriefing. Some equated leadership with hierarchy, while others saw it as a “natural” skill. Interestingly, our findings suggest that the notion of leadership is subjective and lacks consensus in the medical field. However, the idea of a “natural leader” can reinforce hierarchical constructs and encourage dominant individuals to take over, which can hinder team performance and psychological safety (Adams & Anantatmula, 2010; Gamble & Christensen, 2022). Although limited research exists on what constitutes a leader in medical debriefing, our study sheds light on the importance of addressing leadership and hierarchy in promoting effective teamwork and creating a culture of safety.

Dominance can be a problem if an individual perceives themselves as someone who is not a natural leader and, therefore, does not want to speak up about an adverse event or discuss something they were not satisfied with (Adams & Anantatmula, 2010). Based on hierarchical bias (Vanstone & Grierson, 2022), some interviewees perceived senior doctors, the surgeon, or the anaesthetist as the primary leaders in the operating room, and as such, considered them to be “natural leaders.” This view reinforces the traditional power hierarchy in healthcare settings, which can impede effective teamwork and hinder psychological safety. Moreover, many interviewees felt that anyone within the multidisciplinary operative team should be able to lead an operative team to debrief. Responses suggest that an individual’s experience and communication skills may be more relevant to a leadership position than their title or speciality. Therefore, the leadership role can be flexible and negotiated. Ideas that those who take the debrief “should” have leadership skills, however, is not necessarily true. Research suggests that those who are perceived to be leaders due to their hierarchical standing may not necessarily be capable of leading teams due to insufficient training and are therefore no more capable than anyone else at taking a debriefing (Greer et al., 2017).

Understanding leadership leads to different ideas of what leadership should be in a medical context. Although there are different understandings of who should be in charge, considerable research indicates that leadership is entirely necessary when used correctly (Berenholtz et al., 2009; Brennan & Davidson, 2019; Sammer et al., 2010). Leadership aims to achieve a collective purpose (A. Edmondson, 2008; Vanstone & Grierson, 2022), and can be a crucial factor in achieving safety and promoting cultural change in various industries (Berenholtz et al., 2009; Brennan & Davidson, 2019; A. C. Edmondson, 2019; McClintock & Fainstad, 2022). To break down hierarchical barriers and promote effective teamwork, it may be useful to consider transformational leadership, collectivistic leadership, and flat hierarchies. Transformational leadership involves leaders engaging with individuals to achieve common goals (Matheson et al., 2020; Smith, 2015). The purpose of transformational leadership is to elevate, inspire, and ensure individuals become more active in themselves (Matheson et al., 2020; Smith, 2015). Transformational leadership is an empirically supported approach understood as a relationship of mutual stimulation and elevation that raises the level of aspirations of both the leader and those led, thereby transforming both (Matheson et al., 2020; Smith, 2015). Research indicates that transformational leadership in healthcare settings has favourable outcomes with individuals realising their capabilities to reach higher levels of performance and personal meaning (Matheson et al., 2020; Smith, 2015).

Another aspect of leadership is a collective approach, where roles and responsibilities are shared, distributed, or rotated amongst team members (Lv & Zhang, 2017; West et al., 2014). By distributing leadership across teams, formal and informal leaders work together to generate actions, promoting involvement in decision-making and a sense of belonging within a group (Lv & Zhang, 2017; West et al., 2014). Flat hierarchies are gaining popularity in healthcare, as they provide the flexibility and equality needed in a caring environment where all individuals should feel safe to raise concerns and voice opinions (Green et al., 2017). With flat hierarchies, employees have more responsibility for each staff member, as there are more people available for support and guidance (Green et al., 2017). Despite various leadership approaches introduced to healthcare, there is no “one size fits all.”

### Space-time

One of the primary obstacles to debriefing was finding a suitable time and place for the whole team to meet and discuss. Participants demonstrated a trade-off between idealism and pragmatism when deciding when to debrief. According to Kivetz and Tyler (2007), people shift between these positions based on their timeframe. A distal time perspective focuses on the core aspects of the self, allowing for the expression of the idealistic self. In contrast, a proximal time perspective directs attention towards situational contingencies that are incidental to one’s true self, leading to the activation of the pragmatic self (Kivetz & Tyler, 2007). When it comes to debriefing, theatre staff may prefer an idealistic approach if it happens in the future, but a more pragmatic approach if it needs to occur immediately. Our observations of debriefs confirmed this trade-off between the ideal and practical. Debriefing after the patient left for recovery and all the staff returned to the room appeared to be better structured, with fewer encumbrances and distractions, than debriefs that occurred while the patient was still present. However, as noted in several interviews, getting everyone back into the theatre after the last patient left was unlikely for most operating lists.

### Learning to Debrief

Debriefing is a vital component of critical event management in healthcare settings, requiring specific skills for effective implementation (Abbott et al., 2018); however, as expressed by one participant, “no-one is born knowing how to do this”. As Abbott et al. (2018) note, no one is born with an innate ability to debrief, highlighting the need for training and modelling of debriefing skills. From a social constructionist perspective, debriefing skills are acquired through training, education, and modelling from experienced professionals who provide feedback and guidance (Adams & Anantatmula, 2010). Consequently, debriefing skills can be enhanced through practice and reflection with support from colleagues and mentors.

Therefore, debriefing training and education should be included in healthcare organisations’ continuous professional development programs to enhance the quality and safety of patient care. Healthcare professionals must have access to ongoing training and education to develop and improve their debriefing skills. The development of a culture of routine debriefs and standardised debriefing programs can facilitate the acquisition and improvement of debriefing skills and ultimately improve patient safety in healthcare settings.

### Limitations

Debriefing had been performed on a relatively ad hoc basis in our operating rooms, so interviewees had varying degrees of experience. Further research on the experience of operating room workers with regular debriefing would be of value. The study was set in a tertiary paediatric hospital. Subsequent research across a range of hospitals and specialties could provide further insights. Voluntary participation in this study could introduce sampling bias. Although the results of this research were shared with the theatre unit where the research took place, formal reviewing of the themes with interviewees (member checking) was not performed. In keeping with the critical realist paradigm, some of the themes were more descriptive while others were developed (in the discussion) along a more constructionist (critical) line; however, we believe this will enable readers to reflect on some of the practical issues as well as the social issues around debriefing.

## Conclusion

In conclusion, this study of the insights and experiences of frontline workers in the operating room shows that regular, structured debriefs could improve psychological safety, help counter the negative effects of hierarchies and power structures, and promote teamwork. Debriefs were seen as a vehicle for individual and team learning, with the potential to contribute to organisational change. However, to set up an effective debriefing program, it is essential to create a psychologically safe working environment where individuals can be open and confident in group situations without fear of retribution. Creating trust by incorporating programs that acknowledge positive recognition for reporting is crucial to building trust and establishing that reporting will not negatively affect the staff member voicing their concerns or the member of staff involved in a report. Hierarchies can impede the implementation of just culture and psychological safety. Therefore, it is crucial to recognise and address the negative effects of power hierarchies in the workplace. By doing so, healthcare professionals can foster an environment that values open communication, teamwork, and safety, ultimately leading to improved patient care and safety.

## Data Availability

All data produced in the present study are available upon reasonable request to the authors

## Acknowledgements

The authors sincerely thank Dr. Haare Williams for his expert advice and support.

---

**Figure S1.**
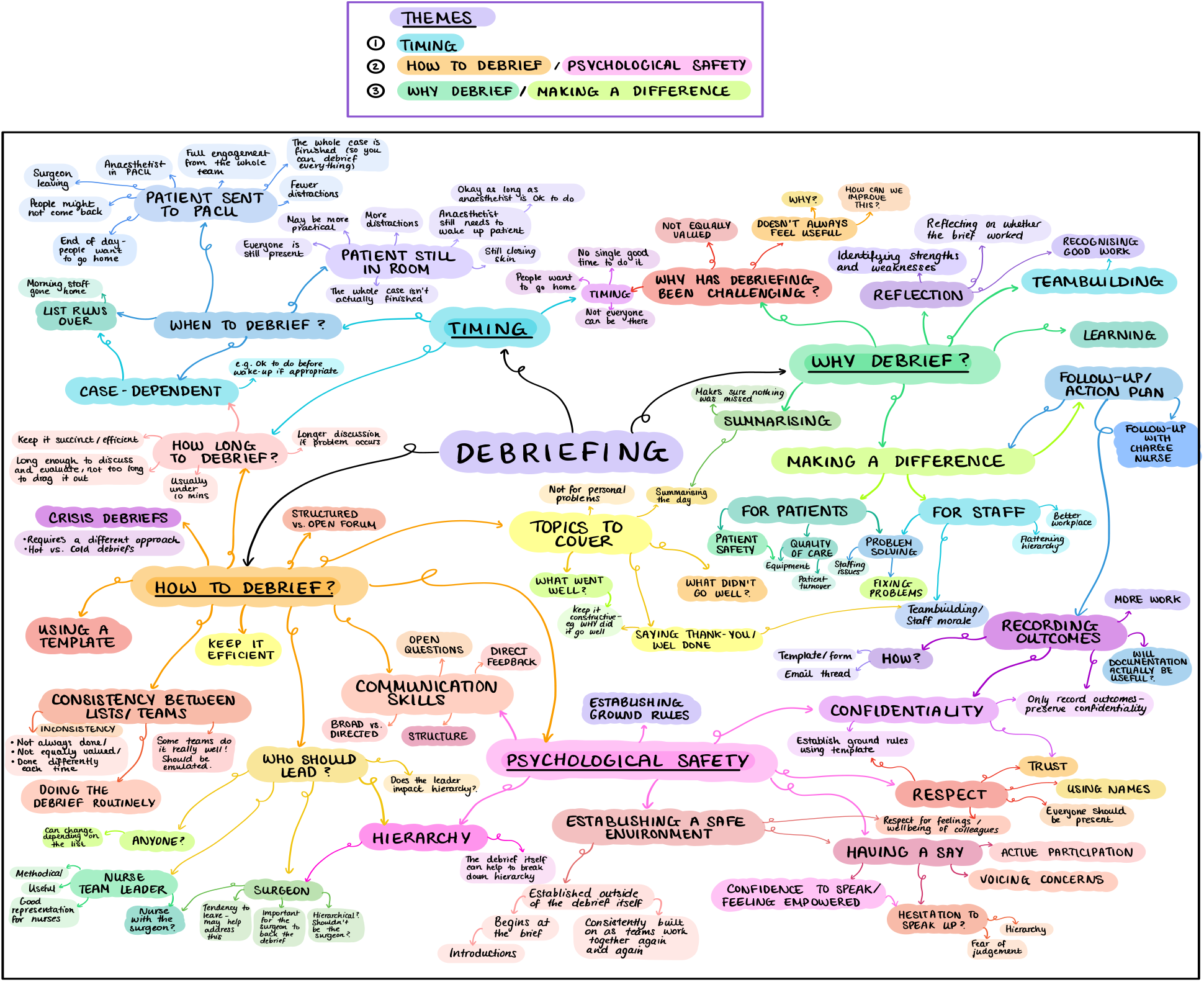
Initial themes map.

