## Appendix 1 for "Psychological safety, hierarchy, and other issues in operating room debriefing: reflexive thematic analysis of interviews from the frontline"

### Interview guide

#### Notes for the interviewer

Start the interview by establishing connection, telling a little about yourself and finding out something about your interviewee. Try to gather all the demographic information. Briefing and debriefing questions are a general guide—they do not all have to be covered. Ask questions appropriate to the focus of your interview. Open-ended questions are good. Prompts are useful— “Can you tell me more about this?”, “What would that look like?” It is a conversation. Let them tell their story.

#### Introduction

Thank you for agreeing to participate in this interview. I am interviewing you to find out what theatre staff think about briefing and debriefing and how we can improve the way we do this. There are no right or wrong answers to any questions, we are interested in your experiences, thoughts and ideas.

Participation in this interview is voluntary. The interview will probably take about 30 – 60 minutes. With your permission I would like to record this interview so as to not miss any comments. The recording will be transcribed and kept confidential. After I have transcribed it I will send you a copy so that you can check it and add/correct anything you like. Any information from your interview that appears in our report will not identify you. You may decline the interview or stop at any time.

Do you have any questions? Let’s start the interview.

##### Informed consent

Explanation—information sheet (if performed on teleconferencing, email information sheet and consent form before the interview)

Consent form signed (if performed on teleconferencing, ask participant to email concert form back)

##### Collect basic demographic information

Have you had a chance to fill out the survey yet?

How long have you worked in operating rooms – years

##### Debriefing topics to cover

Why debrief?

Who should/should not be present at debriefing?

Who should lead debriefs?

What should/should not be debriefed?

What phases of the debrief are important—when and why?

What sort of communication/questioning skills or techniques are useful in debriefs?

Where should debriefing take place?

When should debriefing occur?

How should we set up a safe learning environment at the start of each debrief and ensure psychological safety for everyone?

Debriefings are like a kōrerorero or discussion—how can we ensure everyone has their say?

How long is appropriate?

How should debriefing outcomes be recorded?

For critical incidents, what do you think about debriefing at a later time (when everything has cooled off) and how and when could this be done?

#### Briefings topics to cover

Have you experienced a briefing with a karakia and what was your experience?

What do you think about Māori cultural values in the briefing and in theatre in general?

Briefings are like a kōrerorero or discussion—how can we ensure everyone has their say?

What effect could a karakia have on people's awareness of environmental / sustainability issues? Should we do one for debriefings too?

#### Wrap up

Anything else they would like to add

Summary

Thanks
