## Appendix 2 for "Psychological safety, hierarchy, and other issues in operating room debriefing: reflexive thematic analysis of interviews from the frontline"

### Quality checklist

Braun and Clarke's 15-point checklist of criteria for good thematic analysis (Braun & Clarke, 2006) and our quality self-assessment.

| | Criteria | Our assessment |
|---|---|---|
| 1 | The data have been transcribed to an appropriate level of detail, and the transcripts have been checked against the tapes for 'accuracy'. | Transcripts were transcribed in full except for one interview in which recording failed. Participants were given the opportunity to review and correct transcripts. |
| 2 | Each data item has been given equal attention in the coding process. | Yes. |
| 3 | Themes have not been generated from a few vivid examples (an anecdotal approach), but instead the coding process has been thorough, inclusive and comprehensive. | Themes were developed from examples across the dataset after thorough review and re-review of data. |
| 4 | All relevant extracts for all each theme have been collated. | Collated in Taguette®. |
| 5 | Themes have been checked against each other and back to the original data set. | Yes. |
| 6 | Themes are internally coherent, consistent, and distinctive. | We believe so. |
| 7 | Data have been analysed − interpreted, made sense of − rather than just paraphrased or described. | We have taken a critical approach in analysis and interpretation. |
| 8 | Analysis and data match each other − the extracts illustrate the analytic claims. | Yes. |
| 9 | Analysis tells a convincing and well-organized story about the data and topic. | We believe so. |
| 10 | A good balance between analytic narrative and illustrative extracts is provided. | We believe the balance is about right. |
| 11 | Enough time has been allocated to complete all phases of the analysis adequately, without rushing a phase or giving it a once-over-lightly. | We have reflected and analysed data over a period of about nine months. |
| 12 | The assumptions about, and specific approach to, thematic analysis are clearly explicated. | Yes (see the methodology section). |
| 13 | There is a good fit between what you claim you do, and what you show you have done − i.e., described method and reported analysis are consistent. | We believe so. |
| 14 | The language and concepts used in the report are consistent with the epistemological position of the analysis. | We believe so. |
| 15 | The researcher is positioned as active in the research process; themes do not just 'emerge'. | Researcher reflexivity is stated in the methodology section, and the researchers have actively developed the themes. |

Braun, V., & Clarke, V. (2006). Using thematic analysis in psychology. *Qualitative Research in Psychology*, *3*(2), 77–101. https://doi.org/10.1191/1478088706qp063oa
