## Appendix 3 for "Psychological safety, hierarchy, and other issues in operating room debriefing: reflexive thematic analysis of interviews from the frontline"

### Conceptual depth scale

Nelson's 5-point conceptual depth scale (Nelson, 2017) and our quality self-assessment.

| Criteria | Our assessment |
|---|---|
| Range | A wide range of codes.<br>Data source is quite uniform (interviews of operating room staff). |
| Complexity | Quite a complex initial coding tree as shown in the figures. |
| Subtlety | We have taken a rich interpretation of the language used by participants particularly for the themes on psychological safety and leadership. |
| Resonance | We have engaged extensively with the literature. |
| Validity | We have not performed interviews with staff from other hospitals to check the 'external' applicability of themes. |

Nelson, J. (2017). Using conceptual depth criteria: Addressing the challenge of reaching saturation in qualitative research. *Qualitative Research*, *17*(5), 554–570. https://doi.org/10.1177/1468794116679873
