## Appendix 4 for "Psychological safety, hierarchy, and other issues in operating room debriefing: reflexive thematic analysis of interviews from the frontline"

### Quality reporting guidelines

Braun and Clarke's (2021) tool for evaluating thematic analysis (TA) manuscripts for publication to guide the assessment of TA research quality.

| | Adequate choice and explanation of methods and methodology | Location in the manuscript/explanation |
|---|---|---|
| 1 | Do the authors explain why they are using TA, even if only briefly? | Qualitative approach and research paradigm |
| 2 | Do the authors clearly specify and justify which type of TA they are using? | Qualitative approach and research paradigm |
| 3 | Is the use and justification of the specific type of TA consistent with the research questions or aims? | Qualitative approach and research paradigm |
| 4 | Is there a good 'fit' between the theoretical and conceptual underpinnings of the research and the specific type of TA (i.e. is there conceptual coherence)? | Qualitative approach and research paradigm |
| 5 | Is there a good 'fit' between the methods of data collection and the specific type of TA? | Qualitative approach and research paradigm/Data collection |
| 6 | Is the specified type of TA consistently enacted throughout the paper? | Yes |
| 7 | Is there evidence of problematic assumptions about, and practices around, TA? | No |
| 8 | Are any supplementary procedures or methods justified, and necessary, or could the same results have been achieved simply by using TA more effectively? | N/A |
| 9 | Are the theoretical underpinnings of the use of TA clearly specified (e.g. ontological, epistemological assumptions, guiding theoretical framework(s)), even when using TA inductively (inductive TA does not equate to analysis in a theoretical vacuum)? | Qualitative approach and research paradigm |
| 10 | Do the researchers strive to 'own their perspectives' (even if only very briefly), their personal and social standpoint and positioning? | Researcher characteristics and reflexivity |
| 11 | Are the analytic procedures used clearly outlined, and described in terms of what the authors actually did, rather than generic procedures? | Data processing & analysis |
| 12 | Is there evidence of conceptual and procedural confusion? For example, reflexive TA (e.g. Braun and Clarke 2006) is the claimed approach but different procedures are outlined such as the use of a codebook or coding frame, multiple independent coders and consensus coding, inter-rater reliability measures, and/or themes are conceptualised as analytic inputs rather than outputs and therefore the analysis progresses from theme identification to coding (rather than coding to theme development). | No |
| 13 | Do the authors demonstrate full and coherent understanding of their claimed approach to TA? | We hope so, but are always learning |
| | A well-developed and justified analysis | |
| 14 | Is it clear what and where the themes are in the report? | Figure 1 and Table 2 |

| | | |
|---|---|---|
| | Would the manuscript benefit from some kind of overview of the analysis: listing of themes, narrative overview, table of themes, thematic map? | |
| 15 | Are the reported themes topic summaries, rather than 'fully realised themes' – patterns of shared meaning underpinned by a central organising concept?<br>● If so, are topic summaries appropriate to the purpose of the research?<br>○ If the authors are using reflexive TA, is this modification in the conceptualisation of themes explained and justified?<br>● Have the data collection questions been used as themes?<br>● Would the manuscript benefit from further analysis being undertaken, with the reporting of fully realised themes?<br>● Or, if the authors are claiming to use reflexive TA, would the manuscript benefit from claiming to use a different type of TA (e.g. coding reliability or codebook)? | No, although the theme, "one of the most valuable things" came from inductive coding, it was clearly a strong theme that people really valued debriefing, and this fitted with our critical realist paradigm. |
| 16 | Is non-thematic contextualising information presented as a theme? (e.g. the first 'theme' is a topic summary providing contextualising information, but the rest of the themes reported are fully realised themes). If so, would the manuscript benefit from this being presented as non-thematic contextualising information? | No |
| 17 | In applied research, do the reported themes have the potential to give rise to actionable outcomes? | Conclusions |
| 18 | Are there conceptual clashes and confusion in the paper? (e.g. claiming a social constructionist approach while also expressing concern for positivist notions of coding reliability, or claiming a constructionist approach while treating participants' language as a transparent reflection of their experiences and behaviours) | No |
| 19 | Is there evidence of weak or unconvincing analysis, such as: Too many or two few themes? Too many theme levels? Confusion between codes and themes? Mismatch between data extracts and analytic claims? Too few or too many data extracts? Overlap between themes? | We hope that the balance is about right. |
| 20 | Do authors make problematic statements about the lack of generalisability of their results, and or implicitly conceptualise generalisability as statistical probabilistic generalisability? | No |

Braun, V., & Clarke, V. (2021). One size fits all? What counts as quality practice in (reflexive) thematic analysis? *Qualitative Research in Psychology*, *18*(3), 328–352. https://doi.org/10.1080/14780887.2020.1769238
