## Appendix 5 for "Psychological safety, hierarchy, and other issues in operating room debriefing: reflexive thematic analysis of interviews from the frontline"

### Quality reporting guidelines

Standards for Reporting Qualitative Research (O’Brien et al., 2014).

| | Topic | Location /explanation |
|---|---|---|
| 1 | Title | Title “reflexive thematic analysis” |
| 2 | Abstract | Structured abstract |
| 3 | Problem formulation | Introduction |
| 4 | Purpose or research question | Introduction last paragraph |
| 5 | Qualitative approach and research paradigm | Methodology |
| 6 | Researcher characteristics and reflexivity | Methodology |
| 7 | Context | Methodology |
| 8 | Sampling strategy | Methodology |
| 9 | Ethical issues pertaining to human subjects | Methodology (ethics number) |
| 10 | Data collection methods | Methodology |
| 11 | Data collection instruments and technologies | Methodology |
| 12 | Units of study | Results first paragraph and Table 1 |
| 13 | Data processing | Methodology |
| 14 | Data analysis | Methodology |
| 15 | Techniques to enhance trustworthiness | Methodology |
| 16 | Synthesis and interpretation | Results |
| 17 | Links to empirical data | Results, quotations |
| 18 | Integration with prior work, implications, transferability, and contribution(s) to the field | Discussion |
| 19 | Limitations | Limitations section of discussion |
| 20 | Conflicts of interest | Title page |
| 21 | Funding | Title page |

O’Brien, B. C., Harris, I. B., Beckman, T. J., Reed, D. A., & Cook, D. A. (2014). Standards for Reporting Qualitative Research: A Synthesis of Recommendations. *Academic Medicine*, *89*(9), 1245–1251. https://doi.org/10.1097/ACM.0000000000000388
